# A vulnerability analysis for the management of and response to the COVID-19 epidemic in the second most populous state in Brazil

**DOI:** 10.1101/2020.07.20.20158345

**Authors:** Igor Silva Campos, Vinícius Ferreira Aratani, Karina Baltor Cabral, Jean Ezequiel Limongi, Stefan Vilges de Oliveira

## Abstract

The COVID-19 pandemic has the potential to affect all individuals, however in a heterogeneous way. In this sense, identifying specificities of each location is essential to minimize the damage caused by the disease. Therefore, the aim of this research was to assess the vulnerability of the 853 municipalities in the second most populous state in Brazil, Minas Gerais (MG), in order to direct public policies. Then, an epidemiological study was carried out based on Multi-Criteria Decision Analysis (MCDA) using indicators with some relation to the process of illness and death caused by COVID-19. The indicators were selected by a literature review and categorized into: demographic, social, economic, health infrastructure, population at risk and epidemiological. The variables were collected in Brazilian government databases at the municipal level and evaluated according to MCDA, through the Program to Support Decision Making based on indicators (PRADIN). Based on this approach, the study performed simulations by category of indicators and a general simulation that allowed to divide the municipalities into groups of 1 to 5, with 1 being the least vulnerable and 5 being the most vulnerable. The groupings of municipalities were exposed in their respective mesoregions of MG in a thematic map, using the software Tabwin 32. The results revealed that the mesoregion of Norte de Minas stands out with more than 40% of its municipalities belonging to group 5, according to economic, social and health infrastructure indicators. Similarly, the Jequitinhonha mesoregion exhibited almost 60% of the municipalities in this group for economic and health infrastructure indicators. For demographic and epidemiological criteria, the Metropolitana de Belo Horizonte mesoregion is the most vulnerable, with 42.9% and 26.7% of the municipalities in group 5, respectively. Considering the presence of a population at risk, Zona da Mata reported 42.3% of the municipalities in the most vulnerable group. In the joint analysis of data, the Jequitinhonha, Vale do Mucuri and Vale do Rio Doce mesoregions were the most vulnerable in the state of MG. Thus, through the outlined profile, the present study proved how socioeconomic diversity affects the vulnerability of the municipalities to face COVID-19 outbreak, highlighting the need for interventions directed to each reality.

## Introduction

In late December 2019, hospitals in Wuhan, China, identified numerous patients with pneumonia of unknown cause (1). After investigating the possible etiologic agent involved, on January 7, 2020, Chinese scientists isolated a new type of coronavirus from an individual and, therefore, were able to sequence its genome (2).

The SARS-CoV-2 or 2019-nCoV virus, as it has been named, is the causative agent of the clinical syndrome known as COVID-19 (Coronavirus 19 disease) (3). Although SARS-CoV-2 belongs to the same gender as the viruses responsible for Severe Acute Respiratory Syndrome (SARS) and Middle East Respiratory Syndrome (MERS), the new coronavirus appears to be related to mild infections disorders but with a high rate of transmissibility (3–5). Considering the high levels of transmission, on March 11, the World Health Organization (WHO) characterized COVID-19 as a pandemic due to the rapid spread across countries including Italy, Spain and, later, United States, that currently has the highest number of cases of the novel coronavirus disease (6).

In Brazil, on February 26, 2020, the first case of COVID-19 was confirmed in the state of São Paulo and the first death on March 17, in the same state. During the months of April, May and June, the number of cases and deaths increased exponentially and, then, on June 20, 2020, Brazil was the second country in the world with the highest number of confirmed cases, more than 1 million, and also the second country with the most confirmed deaths, about 50 thousand (6).

In this context, the state of Minas Gerais, the second most populous state in the country, initially stood out for presenting an apparently more controlled situation. While neighboring states in the southeastern region accumulated more than 300,000 cases and almost 22,000 deaths by COVID-19, Minas Gerais was one of the states with the least confirmed cases, approximately 27,000 (6). However, recent researches pointed to a possible underreporting scenario owing to the unprecedented increase in deaths from causes clinically similar to COVID-19, such as SARS, respiratory failure and pneumonia, and due to the low number of tests carried out by the state in comparison with the others, according to data obtained through the Minas Gerais Department of Health (7).

From this perspective, the high number of cases in neighboring states and underreporting in Minas Gerais has rendered the region extremely susceptible to the increase in the number of cases of the disease, with the occurrence and diffusion of new cases. In this situation, delimiting and defining the main regions of vulnerability in the state becomes essential in order to guide the population, managers, public policies and government healthcare workforces. Vulnerability, in this case, relates the chance of exposure to the illness as a result of a set of aspects, both individual and collective (8). Thus, it should be noted that a complex set of indicators may determine a higher or lower vulnerability (9).

In this work, the indicators, selected based on the scientific literature to identify the vulnerability of Minas Gerais cities, trace physical, social and individual characteristics that enable to assess and qualify those regions that have greater difficulty in managing the pandemic of COVID-19. These indicators, divided into demographic, social, economic, health infrastructure, population at risk and epidemiological, are directly related to the increase in illness and death due to the life-threatening condition.

Therefore, the aim of this study was to identify areas of higher vulnerability in the state of Minas Gerais and, from there, assess the region in a segmented and directed way (mesoregions of Minas Gerais) in order to contribute to the elaboration of public policies for prevention and combat of COVID-19.

## Methods

### Study design

This is an epidemiological study to assess vulnerability based on Multi-criteria Decision Analysis (MCDA) (10). Then, indicators were selected in order to allow the assessment of the state of Minas Gerais according to its vulnerability to COVID-19 at the municipal level.

### Study area

The study assesses the state of Minas Gerais (MG), one of the 27 federative units in Brazil, located in the southeastern region of the country, and characterized as the second most populous state and the fourth with the largest territorial extension. It borders the states of São Paulo (south and southwest), Mato Grosso do Sul (west), Goiás (northwest), Bahia (north and northeast), Espírito Santo (east) and Rio de Janeiro (southeast). According to data provided by the Brazilian Institute of Geography and Statistics (IBGE), MG has an estimated population of 21,168,791, demographic density of 33.41 inhabitants per km^2^ (12) in a territory of 586,521,123 km^2^, which corresponds, approximately, to the sum of the territorial areas of Spain and Portugal. The population is predominantly urban, about 85.3% (12), composed of 22.3% of people from 0 to 15 years old, 69.3% from 15 to 64 years old and 8.1% above 65 years old. MG is the state with the third highest gross domestic product (GDP) in Brazil and has Human Development Index (HDI) of 0.731 (11, 12).

The state of MG, whose capital is located in the municipality of Belo Horizonte, has 853 municipalities distributed in 12 mesoregions named: Noroeste de Minas, Norte de Minas, Jequitinhonha, Vale do Mucuri, Triângulo Mineiro and Alto Paranaíba, Central Mineira, Metropolitana de Belo Horizonte, Vale do Rio Doce, Oeste de Minas, Sul e Sudoeste de Minas, Campos das Vertentes and Zona da Mata. Created in 1990 by IBGE, these mesoregions are guided by regional particularities regarding social and administrative processes (13) (Figure 1).

**Figure 1:**
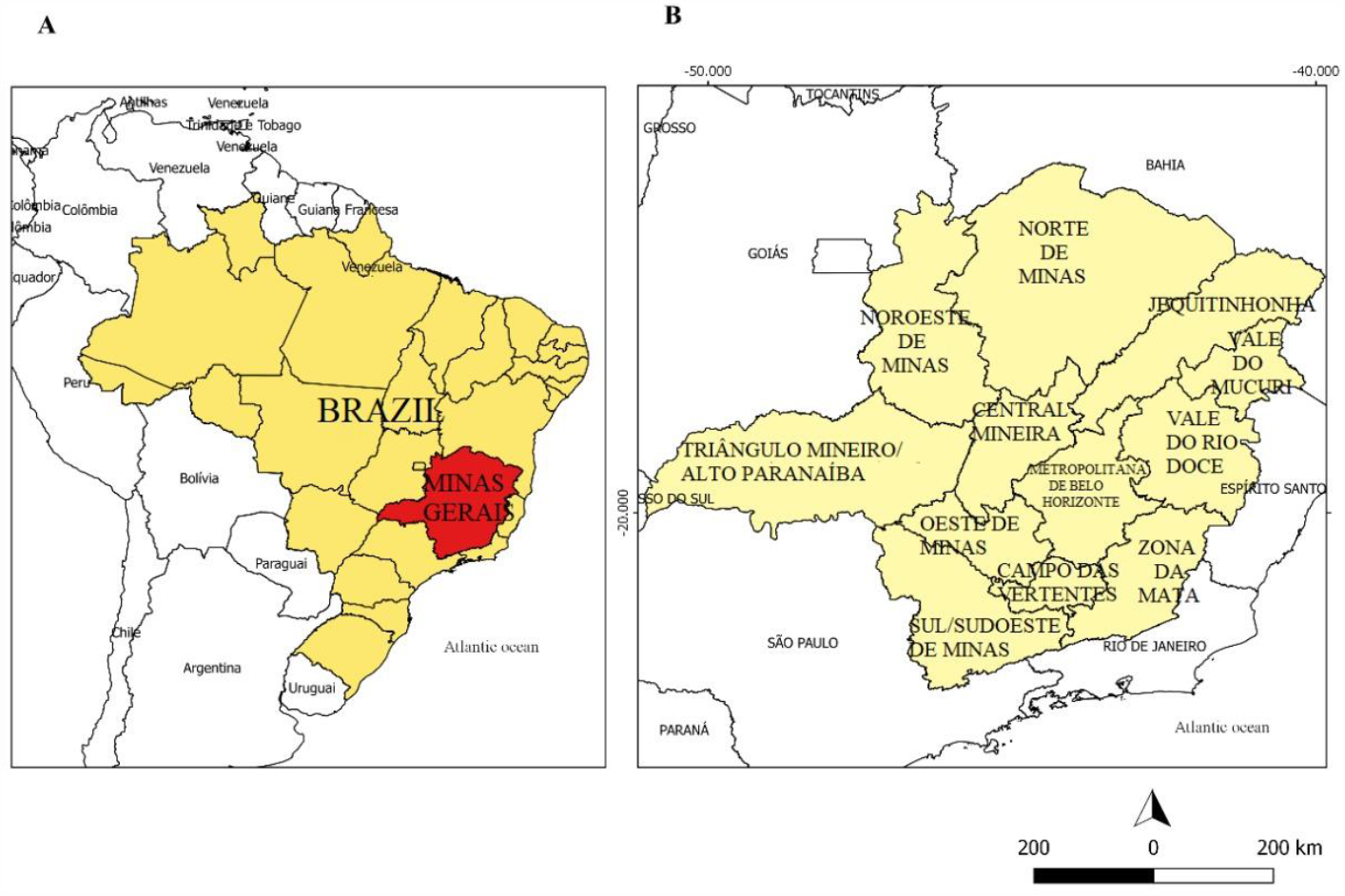
A - Map of Brazil highlighting the state of Minas Gerais in red. B - Map of the state of Minas Gerais divided into its mesoregions.

### Analysis data

The indicators used in this analysis were categorized into: demographic, social, economic, health infrastructure, population at risk and epidemiological. The selection of indicators with some relation to the process of illness and death caused by COVID-19 and their respective categories was conducted based on a literature review in the Scielo and Pubmed databases. Of all the indicators surveyed, those with data related to the municipalities were maintained (Table 1).

**Table 1:** Indicators by category (demographic, social, economic, health infrastructure, population at risk, epidemiological) used in the Multi-criteria Decision Analysis (MCDA) to assess the vulnerability of the municipalities of Minas Gerais to COVID-19.

| Category | Indicators | Description | Application in the context of COVID-19 | Source | References |
| --- | --- | --- | --- | --- | --- |
| **Demographic** | Percentage of the population living in an urban area | Percentage of the population residing in a situation of urban domicile in the municipalities of Minas Gerais. | There is a correlation between the increase in population density, proportion of built area, industrial concentration and other parameters associated with urbanization and increased morbidity by COVID-19. Additionally, air pollution, which is prevalent in locations with high rates of urbanization, contributes to the probability of infections. | 2010 IBGE Population Census | (14,15) |
|  | Demographic density | Demographic density of the territorial unit (Inhabitant per square kilometer) | There is evidence that population density affects the number of COVID-19 daily cases. | 2010 IBGE Population Census | (14, 16,17) |
| **Social** | Percentage of inadequate sanitation | Households that did not present any basic sanitation condition considered appropriate, that is, they were not connected to the general water supply network, to sanitary sewage and had no access to garbage collection. | The virus that causes COVID-19 has already been detected in sewage samples in several countries and in the feces of infected patients, hence demonstrating the need for proper waste treatment. | 2010 IBGE Population Census | (15, 18, 19) |
|  | Illiteracy percentage | Rate of people aged 15 and over who cannot read or write | Important factor of social vulnerability, especially considering that one of the bases for combating the disease is information. Studies also indicate a higher prevalence of certain comorbidities in people with low levels of education. | 2010 IBGE Population Census | (20, 21) |
|  | Gini index | It measures the degree of inequality that exists in the distribution of | Studies indicate that the Gini index can be extremely useful to measure the exposure-disease relationship. | Atlas of Human Development in Brazil 2010 | (22, 23) |
|  |  | individuals according to per capita household income. |  | (http://www.atlasbrasil.org.br/2013/pt/) |  |
|  | Municipal human development index | Geometric mean of the indexes related to income (per capita income indicator), Education (geometric average of the school attendance sub-index, with 2/3 weight, and of the schooling sub-index, with 1/3 weight) and Longevity (obtained through the indicator life expectancy at birth), with equal weights. | The HDI can allow the assessment of social vulnerability by measuring the level of development of each region from three essential factors for quality of life. | Atlas of Human Development in Brazil 2010<br>(http://www.atlasbrasil.org.br/2013/pt/) | (24) |
| **Economics** | Percentage of the population with per capita monthly income | Population considered extremely poor. | Poverty and unemployment, characteristics of a population with such a low monthly income, are social | 2010 IBGE Population Census | (17) |
| | of up to 70 reais (BRL)<br>(equivalent US\$ 13) | | determinants directly related to higher<br>mortality due to COVID-19. | | |
|  | Percentage of the<br>population with health<br>insurance | Percentage of people by<br>municipality who have<br>access to health<br>insurance. | Considering the need to treat more<br>severe cases of the disease in<br>Intensive Care Units (ICUs) and the<br>low availability of beds due to high<br>demand, access to a private health<br>network becomes an important<br>indicator of less vulnerability. | National<br>Supplementary<br>Health Agency<br>TabNet DataSUS<br>(March / 2020) | (25, 26) |
|  | Gross domestic product<br>per capita | Proportion between the<br>wealth produced by a<br>municipality and its<br>number of inhabitants. | The concentration of financial<br>resources facilitates the promotion of<br>measures to contain the pandemic,<br>such as increasing the number of tests<br>in the population. | 2010 IBGE<br>Population<br>Census | (19) |
| **'Health**<br>**infrastructure** | Number of beds per<br>1000 inhabitants | Proportion of the<br>number of<br>hospitalization beds<br>available by<br>municipality. | Due to the pandemic, a great increase<br>in the demand for health services is<br>expected, thus it is essential to<br>identify the most vulnerable regions<br>and optimize the use of services and | TabNet DataSUS<br>- Brazilian<br>National Registry<br>of Health<br>Facilities | (25, 27) |
|  | Number of respirators per 1000 inhabitants | Proportion of the number of available respirators by municipality. | dimension resources that will be necessary to strengthen the response capacity of the health system regionally and locally. | (NRHE) - Physical Resources - 2019 |  |
|  | Number of doctors per 1000 inhabitants | Proportion of the number of doctors available by municipality. |  | TabNet DataSUS - Brazilian National Registry of Health Establishments (NRHE) - Human Resources - 2019 |  |
|  | Number of nurses per 1000 inhabitants | Proportion of the number of nurses available by municipality. |  |  |  |
|  | Number of rapid tests per 1000 inhabitants | Proportion of the number of rapid tests for COVID-19 carried out by municipality. | Although total population testing is impractical, a well-designed program is essential to determine the prevalence of COVID-19 in the general society, in specific subgroups (including healthcare workers) and at-risk groups. | Data provided by the State Health Department of Minas Gerais on 06/22/2020 | (28) |
|  | Number of molecular tests (RT-PCR) per 1000 inhabitants | Proportion of the number of molecular tests (RT-PCR) |  |  |  |
|  |  | performed by municipality. |  |  |  |
| **Population at risk** | Percentage of the population aged 60 or over | Percentage of the resident population in the municipalities of Minas Gerais aged 60 or over. | According to the World Health Organization, the mortality rate due to COVID-19 increases with older age, with higher mortality among people over 80 years old. | 2010 IBGE Population Census | (29) |
|  | Mortality due to diseases of the respiratory system per 1000 inhabitants | Deaths per residence - Chapter ICD-10: X. Diseases of the respiratory system in 2018. | Cancer, hypertension, diabetes, Chronic Obstructive Pulmonary Disease (COPD), heart and cerebrovascular diseases are major risk factors for patients with COVID-19. Thus, municipalities with numerous cases of these life-threatening conditions become more vulnerable. | TabNet DataSUS Mortality Information System - MIS - 2018 | (29, 30, 31) |
|  | Mortality from diabetes per 1000 inhabitants | Deaths per residence - Chapter ICD-10: IV. Diabetes (E10-E14) in 2018. |  |  |  |
|  | Mortality from neoplasms per 1000 inhabitants | Deaths per residence - Chapter ICD-10: II. Neoplasms (tumors) in 2018. |  |  |  |
|  | Mortality due to diseases of the circulatory system per 1000 inhabitants | Deaths per residence - Chapter ICD-10: IX. Circulatory system diseases in 2018. |  |  |  |
| **Epidemiological** | Incidence of COVID-19 | Proportion between new cases of a municipality's COVID-19 and its population. | These occurrence measures are essential to compose an overview of COVID-19 in the municipalities of Minas Gerais, in addition to informing the evolution of the infectious illness in the state, a fact that would not be achieved only with the exposure of the absolute data of cases and deaths from the disease. | Epidemiological bulletin of the Secretary of Health of Minas Gerais on 06/22/2020 | (31) |
|  | Mortality of COVID-19 | Number of COVID-19 deaths per 1000 inhabitants. |  |  |  |
|  | Lethality of COVID-19 | Proportion between the number of deaths by COVID-19 and the population affected by the disease. |  |  |  |

After selecting the indicators, these variables were collected from the databases of the Brazilian Institute of Geography and Statistics (“IBGE”) (https://www.ibge.gov.br/), Atlas of Human Development of Brazil 2013 (http://www.atlasbrasil.org.br/2013/pt/consulta/), from the National Supplementary Health Agency (NSHA) (http://www.ans.gov.br/), Mortality Information System (MIS) (http://www2.datasus.gov.br/DATASUS/index.php?area=060701), the National Registry of Health Establishments (NRHE) (http://cnes.datasus.gov.br/) and of the State Secretariat of Health of Minas Gerais (https://www.saude.mg.gov.br/). Then, 23 indicators remained, grouped according to categories (Table 1). The tabulated absolute data were adjusted to the respective values in percentage, incidence or mortality per 1000 inhabitants, using information from the population projection of the state of Minas Gerais for the year 2019 (11).

### Statistical Analysis

The vulnerability of the municipalities of Minas Gerais to COVID-19 was evaluated by a complex set of factors that, in the present study, are represented by the indicators (32). For the grouped analysis of these factors, so that they started to represent the vulnerability of the municipalities in Minas Gerais, an instrument known as Multi-criteria Decision Analysis (MCDA) was applied through the Program to Support Decision Making Based on Indicators (PRADIN) software (33).

The use of this approach in Health Surveillance is recommended by the Ministry of Health of Brazil as an analysis methodology for the management process of the Brazilian Unified Health System (SUS) (34). In this sense, the method has already been used to classify areas of vulnerability for *Trypanosoma cruzi* (35) and to map the vulnerability to hantavirus infections in Brazil (36). Likewise, in the United States, the methodology was used to analyze the risk of Chagas disease in Texas (37).

By using the PROMETHEE (Preference Ranking Method for Enrichment Evaluation) method, the MCDA tool associates the selected indicators in a hierarchical way, enabling the construction and comparison of different vulnerability scenarios from which different decisions can be inferred (33). In this work, all indicators received the same weight in the hierarchy process, since in the review carried out, no evidence was found on which factor influences COVID-19’s illness and death processes to a higher or lower extent.

The analysis was performed through simulations that, at first, occurred in a segmented manner, according to groupings, evaluating the group of indicators by the pre-defined categories (Table 2). Posteriorly, an analysis was made from a joint simulation of all multi-criteria indicators in order to establish the general vulnerability scenario for Minas Gerais at the municipal level for COVID-19. Six simulations were conducted, with the groupings of indicators and a general simulation, covering all 23 indicators, therefore totalling seven simulations (Table 2).

**Table 2:** Simulations performed by category of indicators (1–6) and the general simulation (7), gathering all the indicators simultaneously.

| Indicators | Simulations |  |  |  |  |  |  |
| --- | --- | --- | --- | --- | --- | --- | --- |
|  | 1<sup>a</sup> | 2<sup>a</sup> | 3<sup>a</sup> | 4<sup>a</sup> | 5<sup>a</sup> | 6<sup>a</sup> | 7<sup>a</sup> |
| **Demographic** |  |  |  |  |  |  |  |
| Population percentage living in urban area | X |  |  |  |  |  | X |
| Demographic density | X |  |  |  |  |  | X |
| **Social** |  |  |  |  |  |  |  |
| Percentage of inadequate sanitation |  | X |  |  |  |  | X |
| Human Development Index |  | X |  |  |  |  | X |
| Illiteracy percentage |  | X |  |  |  |  | X |
| Gini index |  | X |  |  |  |  | X |
| **Economic** |  |  |  |  |  |  |  |
| Population percentage with monthly income bigger than 70 reais (equivalent US\$ 13) | | | X | | | | X |
| Population percentage with health insurance |  |  | X |  |  |  | X |
| Gross Domestic Product |  |  | X |  |  |  | X |
| **Healthcare infrastructure** |  |  |  |  |  |  |  |
| Number of respirators by a thousand habitants |  |  |  | X |  |  | X |
| Number of beds by a thousand habitants |  |  |  | X |  |  | X |
| Number of nurses by a thousand habitants |  |  |  | X |  |  | X |
| Number of doctors by a thousand habitants |  |  |  | X |  |  | X |
| Number of rapid tests by a thousand habitants |  |  |  | X |  |  | X |
| Number of mollecular tests (RT-PCR) by a thousand habitants |  |  |  | X |  |  | X |
| **Population at risk** |  |  |  |  |  |  |  |
| Mortality due to respiratory diseases by a thousand habitants |  |  |  |  | X |  | X |
| Mortality due to cardiovascular diseases by a thousand habitants |  |  |  |  | X |  | X |
| Mortality due to neoplasm by a thousand habitants |  |  |  |  | X |  | X |
| Mortality due to diabetes by a thousand habitants |  |  |  |  | X |  | X |
| Population percentage with 60 years or more |  |  |  |  | X |  | X |
| **Epidemiological** |  |  |  |  |  |  |  |
| COVID-19 incidence by a thousand habitants |  |  |  |  |  | X | X |
| COVID-19 mortality by a thousand habitants |  |  |  |  |  | X | X |
| COVID-19 lethality |  |  |  |  |  | X | X |
| **All indicators (general)** | X | X | X | X | X | X | X |

After the analysis of the indicators by MCDA, each simulation was classified into quintiles, according to the multi-criteria indicator (MCI) and divided into groups of vulnerability according to municipalities, mesoregions and population size, classified in an ascending order. Groups 1 and 2 were composed by the Minas Gerais municipalities with the least vulnerability, group 3 exhibits moderate vulnerability and groups 4 and 5 were those with the greatest vulnerability.

After the classification and definition of the groups, the codes of the municipalities and the program Tabwin 32 (http://www2.datasus.gov.br/DATASUS/index) were used to construct the vulnerability maps for COVID-2019. In thematic maps, the darker colors of the municipalities represent greater vulnerability, while the lightest colors indicate lower vulnerability. For the organization and evaluation of the collected data, the regionalization of the municipalities of the state of Minas Gerais in Mesoregions was used (Figure 1).

To compose the data analysis, the concept of population size of each municipality was also used (38), categorizing in small municipalities those with up to 25 thousand inhabitants, in medium-sized municipalities those with between 25 and 100 thousand inhabitants and in large municipalities those with more than 100 thousand inhabitants.

## Results

The results obtained by the multi-criteria analysis of the groupings of indicators are depicted in Figure 2, according to the analysis of the 853 municipalities in Minas Gerais (Supplementary material 1). Table 3 reveals the mesoregions of the most and least vulnerable municipalities in the state of Minas Gerais for COVID-19, in accordance with the grouping of indicators used in the multi-criteria analysis of decision.

**Table 3:**
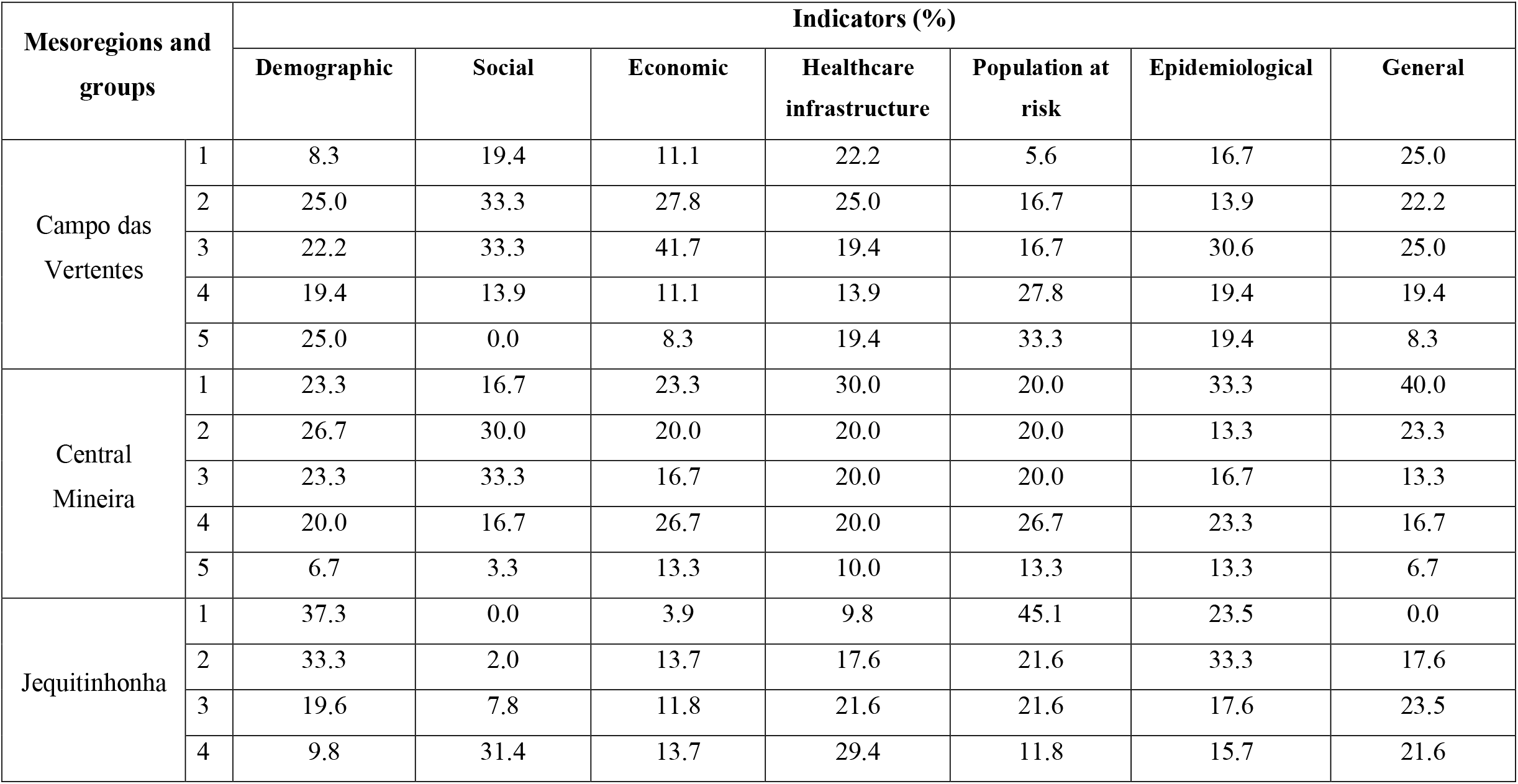
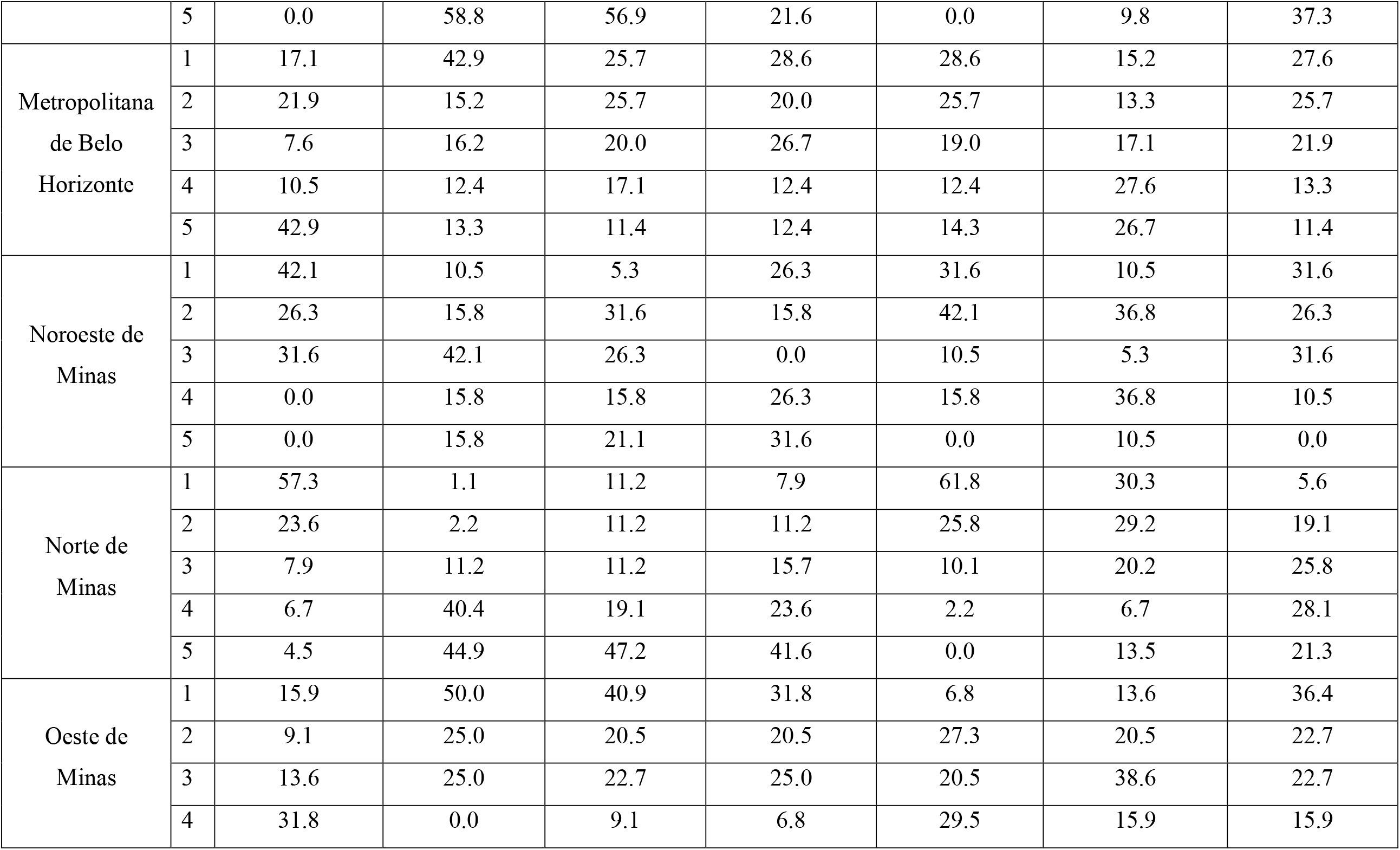
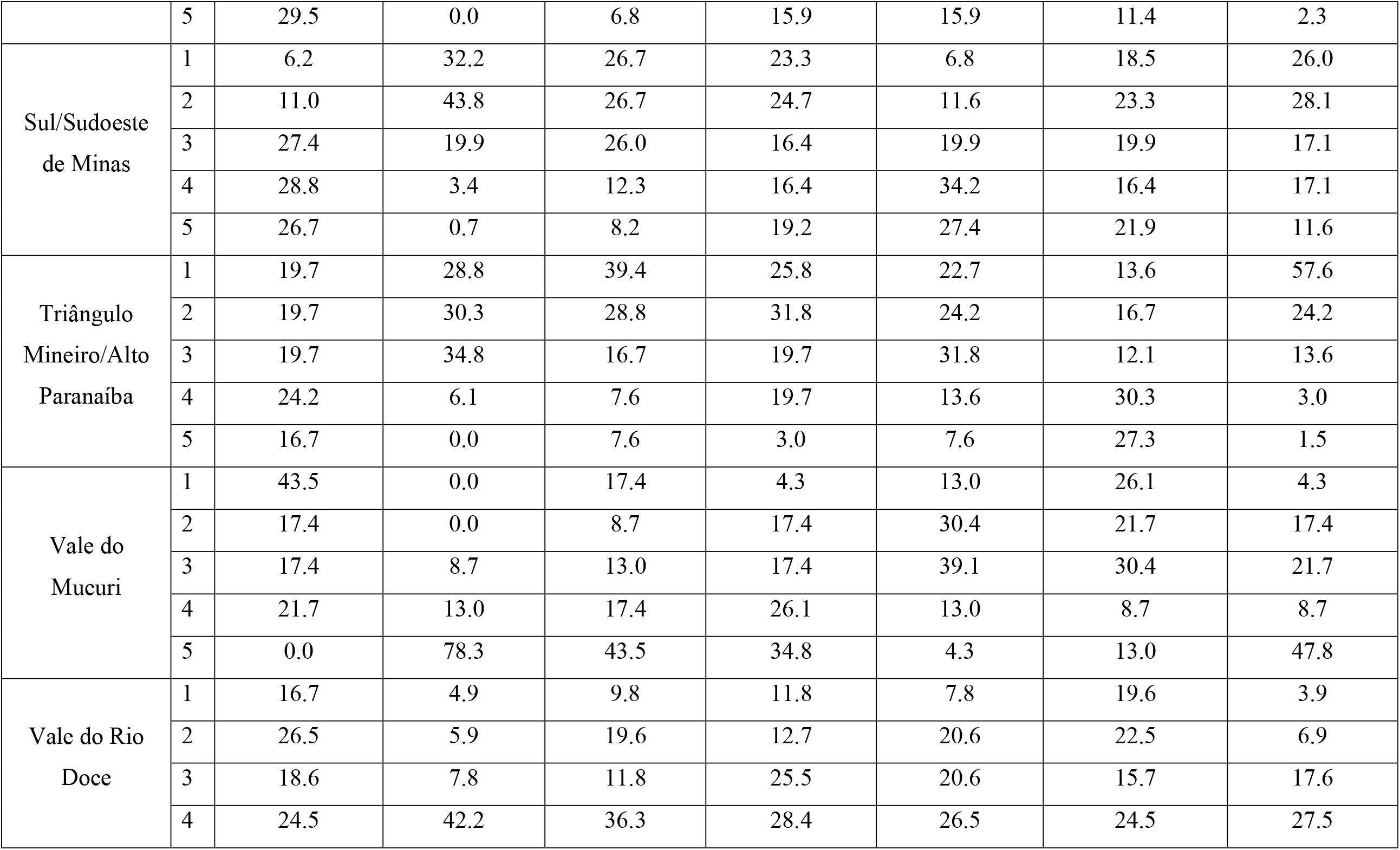
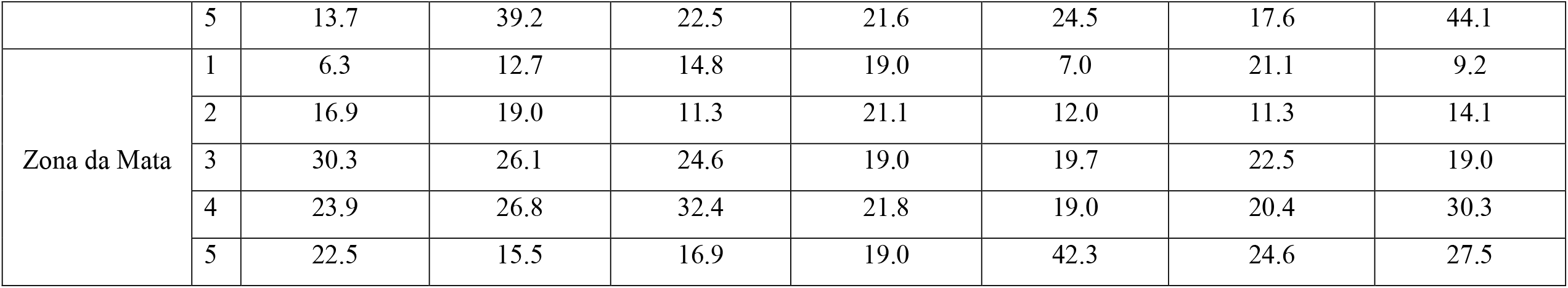
Mesoregions of municipalities and groups of vulnerability (1 and 2 representing lower vulnerability, 3 moderate vulnerability and 4 and 5 greater vulnerability) for COVID-19 in the state of Minas Gerais, according to indicators used in the Multi-criteria Decision Analysis.

**Figure 2:**
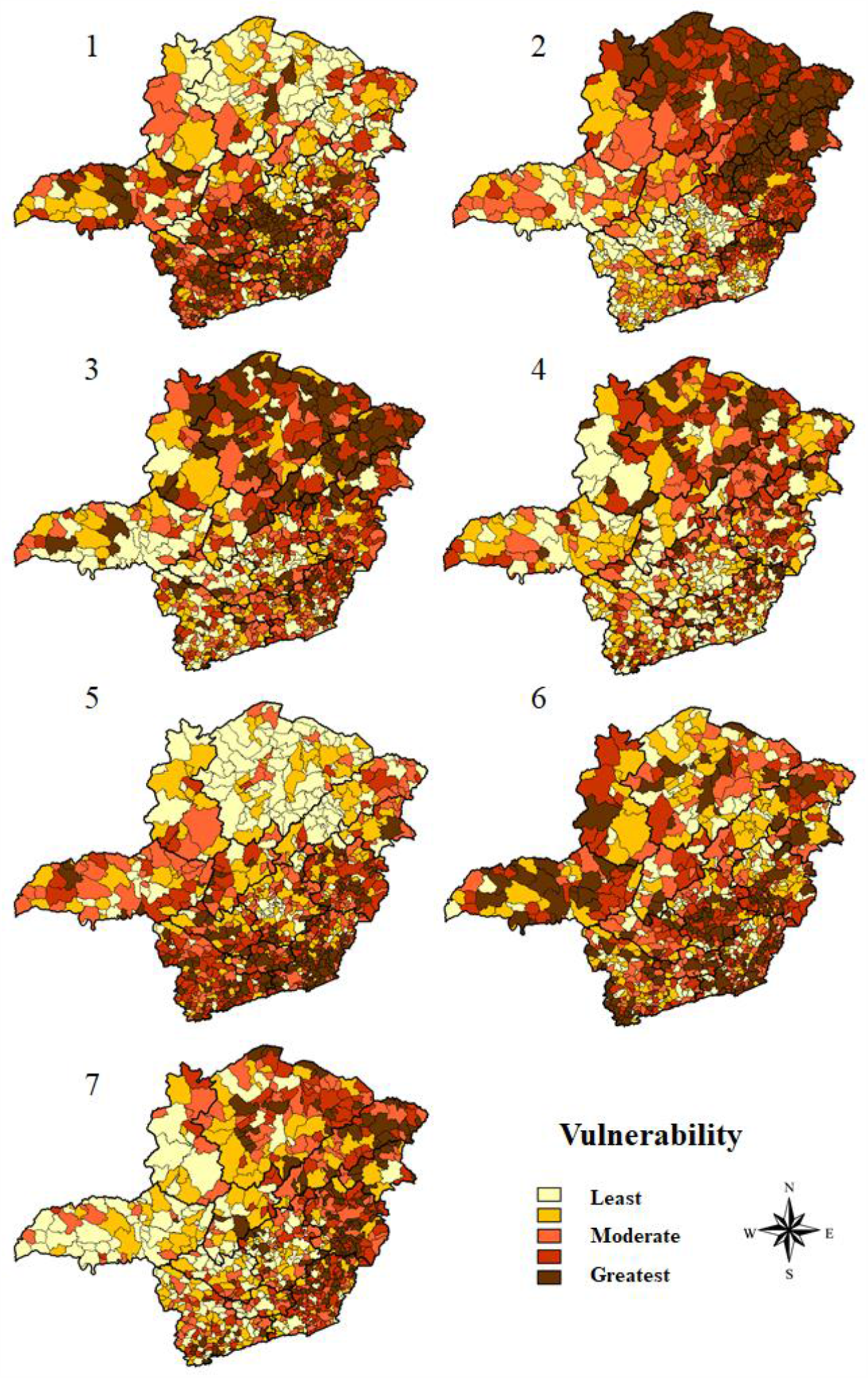
Thematic maps of the vulnerability simulations of the municipalities of Minas Gerais for COVID-19, based on the multi-criteria analysis of decision.

From the analysis of demographic indicators, the results indicated that the Norte de Minas mesoregion is the least vulnerable (Figure 2–1). Accordingly, this mesoregion has 80.9% of the municipalities with the least vulnerability, groups 1 and 2, and 9 municipalities among the 10 least vulnerable in the state for these indicators. Contrarily, the Metropolitana de Belo Horizonte mesoregion had 53.4% of its municipalities classified in groups 4 and 5, that is, with greater vulnerability (Table 3). Thus, 7 of the 10 most vulnerable municipalities in Minas Gerais were in this mesoregion. The findings also revealed that 97.0% of the municipalities categorized as large ones were included in group 5 of vulnerability (Table 4).

**Table 4:** Results of the Multi-criteria Decision Analysis according to the population size of municipalities in the state of Minas Gerais and their vulnerability classification, in accordance with the grouping of indicators used in the multi-criteria analysis of decision for COVID-19.

| Cities by population size | Total of cities | Groups | Indicators (%) |  |  |  |  |  |  |
| --- | --- | --- | --- | --- | --- | --- | --- | --- | --- |
|  |  |  | Demographic | Social | Economic | Healthcare infrastructure (%) | Population at risk | Epidemiological | General |
| Low size<br>(Until 25 thousand habitants) | 705 | 1 | 23.3 | 12.9 | 16.2 | 11.1 | 18.6 | 24.3 | 17.0 |
|  |  | 2 | 22.7 | 19.9 | 20.0 | 20.0 | 18.9 | 21.7 | 18.2 |
|  |  | 3 | 22.8 | 22.3 | 21.7 | 21.3 | 20.6 | 21.8 | 21.1 |
|  |  | 4 | 20.7 | 22.8 | 21.1 | 23.5 | 20.0 | 20.1 | 21.1 |
|  |  | 5 | 10.5 | 22.1 | 21.0 | 24.1 | 22.0 | 12.1 | 22.6 |
| Medium size<br>(25 to 100 mil habitants) | 115 | 1 | 6.1 | 45.2 | 32.2 | 56.5 | 26.1 | 0.0 | 31.3 |
|  |  | 2 | 9.6 | 23.5 | 20.0 | 24.3 | 26.1 | 15.7 | 27.8 |
|  |  | 3 | 8.7 | 11.3 | 13.9 | 13.9 | 16.5 | 14.8 | 14.8 |
|  |  | 4 | 20.9 | 8.7 | 17.4 | 4.3 | 20.9 | 24.3 | 18.3 |
|  |  | 5 | 54.8 | 11.3 | 16.5 | 0.9 | 10.4 | 45.2 | 7.8 |
| Large size<br>(more than 100 thousand habitants) | 33 | 1 | 0.0 | 84.8 | 54.5 | 78.8 | 30.3 | 0.0 | 45.5 |
|  |  | 2 | 0.0 | 12.1 | 21.2 | 6.1 | 24.2 | 0.0 | 33.3 |
|  |  | 3 | 0.0 | 3.0 | 6.1 | 15.2 | 21.2 | 0.0 | 15.2 |
|  |  | 4 | 3.0 | 0.0 | 6.1 | 0.0 | 18.2 | 3.0 | 3.0 |
|  |  | 5 | 97.0 | 0.0 | 12.1 | 0.0 | 6.1 | 97.0 | 3.0 |

From the analysis of social indicators, the results showed that the mesoregions Oeste de Minas and Metropolitana de Belo Horizonte occupy, together, the position of the least vulnerable. Both have 63.0% of their municipalities integrating groups 1 and 2. In contrast, Norte de Minas, Vale do Rio Doce and Vale do Mucuri stand out as the most vulnerable (Figure 2-2). While the first two mesoregions have, respectively, 85.3% and 81.4% of their municipalities distributed between groups 4 and 5, the Vale do Mucuri mesoregion exhibits 91.3% of its municipalities integrating the most vulnerable groups, where 4 of the 10 most vulnerable cities are located in this mesoregion (Table 3). Besides that, a significant part of the large (84.9%) and medium-sized (45.2%) municipalities were among the least vulnerable regarding social indicators (Table 4).

Regarding economic indicators, it was found that Triângulo Mineiro/Alto Paranaíba mesoregion was the least vulnerable region in the state, with 68.2% of the municipalities classified in groups 1 and 2 (Figure 2-3). The Jequitinhonha and Norte de Minas mesoregions had 56.9% and 47.2% of their municipalities categorized in group 5, respectively. Importantly, 6 most vulnerable municipalities in the state belong to these regions (Table 3). In addition, within this analysis category, 52.2% and 75.7% of medium-sized and large cities, respectively, are part of the groups 1 and 2 and, therefore, have lower vulnerability (Table 4).

When analyzing the indicators that assess health infrastructure, the results reported that Oeste de Minas and Central Mineira mesoregions are less vulnerable, with 31.8% and 30.0% of the municipalities in group 1, respectively (Table 3). Norte de Minas and Vale de Mucuri were among the most vulnerable areas, with 41.6% and 34.8% of the municipalities in group 5 (Figure 2-4) (Table 3). In this regard, medium-sized and large municipalities with, respectively, 80.8% and 84.9% in groups 1 and 2, have lower vulnerability (Table 4).

Considering the presence of a risk population, the findings showed that the mesoregion of Norte de Minas was the least vulnerable, with 61.8% of its municipalities classified in group 1. Notwithstanding that, Zona da Mata mesoregion proved to be the most vulnerable, with 42.3% of its municipalities being part of group 5 (Table 3) (Figure 2-5). Furthermore, it is noteworthy that 22.0% of the municipalities classified as small-sized were part of group 5 (Table 4).

Based on epidemiological indicators, the Norte de Minas and Jequitinhonha mesoregions stood out due to their lower vulnerability with, respectively, 59.6% and 56.9% of the municipalities classified in groups 1 and 2. In turn, Metropolitana de Belo Horizonte and Triângulo Mineiro/Alto Paranaíba revealed higher vulnerability, with 54.3% and 57.6% of the municipalities classified in groups 4 and 5, respectively (Table 3) (Figure 2-6). However, 7 of the 10 most vulnerable municipalities in the state are located in the mesoregion of Zona da Mata. Moreover, the findings also reported that 45.2% of medium-sized municipalities and 97.0% of large municipalities were classified in group 5 (Table 4).

The analysis of all indicators jointly demonstrated that Triângulo Mineiro/Alto Paranaíba is the least vulnerable mesoregion (Table 3) (Figure 2-G). With 57.6% of its municipalities classified in group 1, this region contains 5 of the 10 least vulnerable municipalities. It is also possible to highlight the positions of Vale do Mucuri and Vale do Rio Doce. While the first mesoregion has the highest rate of municipalities occupying group 5 (47.8%), the mesoregion of Vale do Rio Doce comprises 5 of the 10 most vulnerable municipalities, with 44.1% of its municipalities integrating group 5 (Table 3) (Figure 2-7). In addition, the general analysis also revealed that small municipalities are among the most vulnerable, with 43.7% of their representatives divided between groups 4 and 5 (Table 4).

## Discussion

The use of MCDA to generate these results was based on the perspective that the health-illness-care process depends on several factors determined by the individuals’ living conditions. Thus, exploring the structure and spatial dynamics of the population is essential for the characterization of health situations to plan actions and allocate resources (15, 39). From this perspective, in order to draw a panorama of reality, the use of indicators becomes an important instrument to measure it in a succinct, objective, quick and efficient way, aiming to support an intervention (39).

In the context of the COVID-19 pandemic, it is emphasized that the novel coronavirus infection has the potential to affect everyone in society, however in a heterogeneous way (40), therefore requiring identification of areas of vulnerability. Although various strategies for mitigating the rate of disease transmission are recommended for the entire community, it is pivotal to examine areas based on their unique characteristics, including demographic variation, economic aspects, health conditions of population and characteristics of the health system, in order to produce improved and targeted interventions (41). Thus, in a state with a projection of more than 20 million inhabitants in 2019 and more than 580 thousand km^2^ of area (IBGE), such peculiarities of each region become even more accentuated.

In this scenario, the state of Minas Gerais occupied, in 2012, the 9th position in the national urbanization ranking (24), but its territorial vastness causes important internal inequalities regarding the urbanization rates of the municipalities, a relevant fact for vulnerability analysis for COVID-19 (42). This is because COVID-19 is closely related with high population density owing to the high degree of social interactions. In this sense, individuals living in urban areas are more likely to test positive for the disease when compared to individuals living in rural areas (43, 44). Thus, regarding the state of Minas Gerais, the lower vulnerability verified in Norte de Minas mesoregion is due to the much lower urbanization rates in comparison to the average state, in addition to the low demographic density and predominance of smaller cities (12). Furthermore, proving this strong relationship, it is important to mention the specificity of Montes Claros (Figure 1-A), a municipality in the Norte de Minas that differs in terms of higher rates of urbanization and, for this reason, exposed a similar profile to the municipalities located in the center-south portion of Minas Gerais, with high vulnerability, comparable to the Metropolitana de Belo Horizonte mesoregion.

According to the Atlas of Social Vulnerability in Brazilian Municipalities (46), by 2015, all thirty municipalities in the Southeast region classified as high social vulnerability were in Minas Gerais. Approaching this reality to the Covid-19 transmission scenario, locations with better education, sanitation and development indicators concentrate greater instructional and sanitary capacity to contain the spread of the virus (18). Thus, in the analysis of vulnerabilities, large and medium-sized municipalities, predominant in the central-southern portion of the state, demonstrated less risk, given that the best social indicators are verified in Oeste de Minas and Metropolitana de Belo Horizonte mesoregions. In contrast, the northern mesoregions, especially Vale do Mucuri, are the ones with the worst educational and housing conditions (46). The northern portion of the state herein proved to be more socially vulnerable to Covid-19, demanding public policies directed to improving these indicators to overcome the contagion.

In addition, from the analysis of economic indicators, the results also indicated greater vulnerability in the northern region of Minas Gerais, mainly in Jequitinhonha and Norte de Minas mesoregions, in contrast to Triângulo Mineiro/Alto Paranaíba and Metropolitana de Belo Horizonte mesoregions. Remarkably, this information is relevant as, in the context of the pandemic, the politicians have focused on populations at risk considering mainly comorbidities and age (42). However, socioeconomic issues have been in the background, which may favor COVID-19 exposure and mortality. Additionally, economically disadvantaged people are more likely to live in accommodation with high number of people and less access to open areas in their homes, besides to having unstable occupations that do not allow remote office work (PATEL et al, 2020). In this sense, the current prevention model based mainly on social isolation can be fragile and limited when applied to needy, isolated and low-educated populations (47). Therefore, poverty represents a hurdle to effective measures to contain the pandemic and must be taken into account in public policy decision making.

Regarding the indicators related to the health infrastructure, a higher vulnerability was also found in the northern regions of the state. This category is especially important when considering more severe cases of the disease, which require hospitalization in Intensive Care Units (ICU) (26). Inadequate health infrastructure directly influences the mortality rate caused by COVID-19. In this context, the Brazilian health regions with the highest mortality rates are located in places where the shortage of ICU beds and ventilators is more prevalent (27). Thus, the saturation of ICUs and respirators resulting from the increasing demand becomes an aggravating factor for the COVID-19 pandemic and requires attention from managers (25). In this context, the construction of temporary hospitals, as has already been done in other parts of Brazil, may be an alternative.

Considering the number of tests for COVID-19, an important factor in determining the prevalence of infection / disease in the population (28), Brazil, as the vast majority of other developing countries, has a very modest number when compared to developed countries (48). Despite being the second country with the highest number of absolute deaths and the fifth with the highest number of deaths per million inhabitants, Brazil is only the fourteenth country testing patients, hence demonstrating a serious concern (49). The context of Minas Gerais is even more worrying, considering that the state has the third lowest number of tests per thousand inhabitants among the 26 states and 1 federative unit in the country (6). This has failed to identify potential transmitters and directly influences the number of reported cases, which may be much smaller than the actual number. In addition, questions have been raised about possible underreporting of cases, which further aggravates the state’s situation, thus making government intervention urgent (7).

The factors of comorbidities and age, which compose the population at risk indicators, were raised during the pandemic in order to draw a well-defined profile of people more susceptible to the complications of COVID-19. Both in Wuhan, China (50) and in the Italian states (51), respiratory and cardiac diseases, as well as neoplasms, diabetes and advanced age are considered, from the beginning, factors for complication of the clinical condition and higher mortality. In this sense, access to health infrastructure and education acts as an aggravation of diseases, and the region that is able to provide more appropriately these resources to the population ensures better conditions to prolong life (52). Importantly, a greater longevity is also accompanied by an increase in the elderly population, which is more affected by chronic diseases, and with regard to COVID-19, act as complicating factors of the clinical condition. Thus, as pointed out by the present study, the worst social and health infrastructure indicators in the Norte de Minas may be associated with lower longevity, hence leading the northern portion of the State to have fewer people in the risk group (53). On the other hand, the Zona da Mata is better assisted, which leads its population to be longer-lived and, consequently, to have a higher number of people more susceptible to the health complications caused by COVID-19.

Another pivotal issue is that coping with the COVID-19 pandemic involves changes in the health system and also requires political decisions that affect the management of chronic non-communicable diseases, as well as patients’ adherence to treatment, especially those from less favored social classes (54). Furthermore, the pandemic scenario increases patients’ fear of seeking health services, which can increase mortality from events related to these chronic illness (55). Thus, based on the identification of this regional vulnerability profile, it is possible to outline public policies that address the major diseases associated with the worsening of the clinical condition of patients with COVID-19, as well as specialized care for the elderly, markedly more affected by the disease.

With regard to epidemiological aspects, greater vulnerability was found in the metropolitan mesoregions of Metropolitana de Belo Horizonte, Triângulo Mineiro/Alto Paranaíba and Zona da Mata and, in a lesser extent, in the Norte de Minas and Vale do Mucuri. In this sense, the observance of the great impact of the mesoregions further south and southeast of the state becomes relevant when considering that these places are located on the border with the states of São Paulo and Rio de Janeiro, which concentrate the highest number of cases in the disease in Brazil. Besides that, the lower socioeconomic development in the North of Minas Gerais favors the scenario of underreporting of cases in the state, which can be associated with a large north-south discrepancy in the numbers found. Thus, social distance should be considered through reliable measures, including travel restrictions or even the institution of lockdown, which have proven effective in countries such as China, South Korea, Iran, Italy, France and the United States (56). In addition to these measures, others widely used worldwide must be promulgated with greater avidity in the most affected municipalities, such as awareness about the use of personal protective equipment, social distance, closing schools, business buildings, quarantine, cleaning and disinfection and increase in the number of tests (19, 57–60).

Considering also the joint analysis of all indicators, the lower vulnerability of the mesoregion of Triângulo Mineiro and Alto Paranaíba was proven, with better social, economic, risk population and health infrastructure indicators. As a result, it is possible to conclude that the highest human development indexes, in addition to a diverse and historically integrated economy in the State of São Paulo, associated with a higher presence of young people and the concentration of hospital resources, integrate factors to reduce vulnerability regarding COVID-19 (61, 62). Similarly, the findings also reported the greatest vulnerability in Vale do Mucuri, followed by Vale do Rio Doce. In these areas, the economy is fragile, basically composed of the primary sector. Education and sanitation indicators are remarkable low and there is a predominance of higher age groups (61). Additionally, the few existing hospital resources are concentrated especially in the municipalities of Teófilo Otoni and Governador Valadares, not reaching all the surrounding municipalities (62).

Besides to the evident differences revealed by the indicators between the mesoregions of the state of Minas Gerais, data also exhibited marked differences between the municipalities grouped by population size, with emphasis on the numerical predominance of small municipalities. Thus, the analysis performed by the present study is essential to better understand the possible particularities of these municipalities in the face of the pandemic, highlighting the need to also formulate specific strategies and public policies according to the size of the population.

The high transmissibility of SARS-COV-2 causes large urban centers with population agglomerations to have a rapid and exponential increase in cases and deaths from the entry of the virus into the population (61). In fact, the rapid increase in the absolute number of cases requires the development of containment measures and, in some cases, when applied with due urgency, relative success is achieved. However, the pandemic is not restricted to large municipalities, but also reaches medium-sized and small areas, consolidating the internalization of the disease, which in Brazil reaches more than 90% of the municipalities (6, 63).

The smaller municipalities did not have large absolute numbers of cases and deaths compared to the others, but when investigating the proportion measures, such as incidence, lethality and mortality, as performed by this work, it is clear that several small municipalities were in a serious situation. In view of this concern, the exposed and disseminated absolute data of these cities do not cause as much impact on the population and public managers as they should, making containment measures such as social distance take time to be employed or adhered to the population and managers.

This scenario is particularly worrying for the small municipalities that were, for the most part, more vulnerable, especially in relation to health infrastructure and financial resources (64). A large portion of these ones reported few ICU beds, few or no respirators and a reduced number of health professionals, hence leading to a high lethality of the disease, since the basic care conditions of the most serious cases of COVID-19 are not guaranteed (41).

Based on the regionalization process defined by the Minas Gerais Health Regionalization Master Plan, these small municipalities should be assisted by medium and large-sized municipalities, having access to medium and high complexity services as needed in severe cases of the disease (27). However, the reality of the state of Minas Gerais does not meet this proposal, as the assessment demonstrated several discontinuities and inequalities in all indicators of the state. Alarmingly, the small municipalities are isolated in the middle of the pandemic, without support from the medium-sized and large municipalities and without enough resources to improve their own health infrastructure (62, 64). Municipalities listed to offer this support face the overcrowding of beds and the lack of respirators (25).

It is important to highlight that some indicators that could contribute to the mapping of the vulnerability of the state of Minas Gerais to COVID-19 were not included due to the absence of data related to the municipalities, hence hindering the tabulation. The data included also had a difference in the dates on which they were made available, since some are only accessed by the 2010 demographic census conducted by IBGE. Another limitation is also related to the database available for consultation, since various indicators may be out of date, especially in small municipalities where the registration process does not occur or is not done properly.

In addition, in the data tabulation process for MCDA, one of the steps consists in defining weights for the different data included, herein determining which ones would have different intensities of influence. However, the present study chose to keep all data with the same weight owing to the absence of evidence, showing which factors would have a higher or lower influence in the pandemic and the difficulty of stipulating the proportion of this influence. Therefore, it is advisable that the maps and the findings of this study are used only as an instrument of guidance for public policies with other existing tools, that is, they should not be used as the only resource. Also, a segmented analysis may be performed by category of indicators in order to avoid possible differences in the influence of indicators in the compilation of the final result.

From the vulnerability analysis carried out, it is clear that the demands of the municipalities of Minas Gerais in the context of COVID-19 are different, varying according to the region in which they are located and their population size. Thus, a public policy planned for the state as a whole will have totally different applicability and effectiveness depending on the region or municipality in question. Therefore, a more segmented analysis of the state should be conducted, as proposed by this work, in order to identify the particularities of each municipality and mesoregion in the search for interventions that have an effect in a faster and more practical way, as the context requires. In this scenario, it is essential to take measures to contain the spread of the disease in the state as a whole, not just in the most economically important regions. For this, the problem of capillarity of the state regarding social, economic and health indicators must be solved, so that everyone has a similar capacity to fight the pandemic. As a result, the state will not only benefit from combating the COVID-19 pandemic, but also from combating all inequalities that have been consolidated in Minas Gerais and directly affect the quality of life of the population in the less-assisted regions.

## Data Availability

All data in this document are in the public domain and the access links have been informed in the text.

